# Demonstrating multi-country calibration of a tuberculosis model using new history matching and emulation package - *hmer*

**DOI:** 10.1101/2022.05.13.22275052

**Authors:** Danny Scarponi, Andrew Iskauskas, Rebecca A Clark, Ian Vernon, Trevelyan J. McKinley, Michael Goldstein, Christinah Mukandavire, Arminder Deol, Chathika Weerasuriya, Roel Bakker, Richard G White, Nicky McCreesh

## Abstract

Infectious disease models are widely used by epidemiologists to improve the understanding of transmission dynamics and disease natural history, and to predict the possible effects of interventions. As the complexity of such models increases, however, it becomes increasingly challenging to robustly calibrate them to empirical data. History matching with emulation is a calibration method that has been successfully applied to such models, but has not been widely used in epidemiology partly due to the lack of available software. To address this issue, we developed a new, user-friendly R package *hmer* to simply and efficiently perform history matching with emulation. In this paper, we demonstrate the first use of *hmer* for calibrating a complex deterministic model for the country-level implementation of tuberculosis vaccines to 115 low-and middle-income countries. The model was fit to 9–13 target measures, by varying 19–22 input parameters. Overall, 105 countries were successfully calibrated. Among the remaining countries, *hmer* visualisation tools, combined with derivative emulation methods, provided strong evidence that the models were misspecified and could not be calibrated to the target ranges. This work shows that *hmer* can be used to simply and rapidly calibrate a complex model to data from over 100 countries, making it a useful addition to the epidemiologist’s calibration tool-kit.

## 1. Introduction

Complex mathematical models, also known as simulators, are widely used in many fields in science and technology, including infectious disease epidemiology. Their utility and reliability in predicting the behaviour of real-world systems strongly depends on our ability to calibrate them to the empirical data. Despite the wide variety of calibration methods available to date, the application of calibration methods to the analysis of complex models is often lacking. A key reason that comprehensive complex model calibration is uncommon is that most formal methods require a vast number of model evaluations, which is exacerbated for high-dimensional input spaces (Kennedy & O’Hagan, 2001). This is often infeasible when dealing with computationally expensive models. The lack of methodologies and tools to robustly calibrate and analyse complex models hinders the credibility and validity of subsequent inferences drawn from model predictions. This can lead to overconfident or misleading recommendations being made to policy makers, potentially costing lives.

History matching with emulation is a robust calibration method that has been applied successfully to complex models in a variety of fields, e.g. cosmology (Vernon et al., 2010), climate science (Williamson et al., 2013), geology (Craig et al., 1996, 1997), and infectious disease epidemiology (Andrianakis et al., 2015; Andrianakis et al., 2017; McKinley et al., 2018). Despite this, its routine use by infectious disease modellers has been hindered by the lack of accessible computer software to implement the various components of the method. Our new R package *hmer* addresses this problem, allowing the user to more easily and efficiently perform history matching with emulation.

We define calibration as identifying all parameter sets that match the observed data, given the included sources of uncertainties. It is important to appreciate that this is slightly different from the technical definition of calibration used in the Uncertainty Quantification literature, where the existence of a single best-fitting parameter set is assumed and calibration is the process of estimating the posterior distribution of the (unknown) best-fitting parameter set.

In this article, we show how we performed history matching with emulation on a complex deterministic model for the implementation of tuberculosis vaccines at country-level, calibrating the model to a total of 105 low-and middle-income countries. Such a large-scale task was made straightforward thanks to our package *hmer*, which allowed us to automate the various steps of the history matching and emulation process. This was possible thanks to inbuilt checks available in *hmer*, which monitored the performance of the process wave by wave.

## 2. The model and the calibration task

Worldwide, tuberculosis was the leading cause of death from a single infectious agent in 2019, killing an estimated 1.4 million people (World Health Organization, 2020). The only licensed vaccine against tuberculosis, Bacillle Calmette-Guerin, is effective at preventing disseminated disease in infants, but confers highly variable efficacy against pulmonary tuberculosis in adults. The development of a new tuberculosis vaccine that protects several categories of people, including adults, the elderly, and immunosuppressed patients, is critical for rapidly reducing disease incidence and mortality.

Contracted by the World Health Organization, the London School of Hygiene and Tropical Medicine Tuberculosis Vaccine Modelling Group developed a model to evaluate the potential impact of country-level implementation of novel tuberculosis vaccines, in order to estimate the broader health and economic impacts (Clark et al., 2022). Full details of the model are provided in Clark et. al, 2022, and described briefly here.

A compartmental deterministic dynamic model of *Mycobacterium tuberculosis* (*Mtb*) transmission and progression was developed, structured by the following core dimensions: 82 age classes, 8 tuberculosis natural history classes, two socio-economic status (SES) classes defined by access-to-care, and, for countries with a moderate or larger contribution of HIV to the tuberculosis epidemic, three HIV and antiretroviral therapy (ART) status classes.

In countries where HIV was not simulated, we had 19 input parameters that were varied during calibration, and 9 calibration targets. Among the 19 parameters, 12 were related to tuberculosis natural history, one to tuberculosis mortality, two to background mortality, three to tuberculosis treatment and one to SES. The 9 calibration targets were all for 2019, and can be divided into the following categories: tuberculosis incidence (all ages, age 0–14, and age 15+), tuberculosis notifications (all ages, age 0–14, and age 15+), tuberculosis mortality (all ages), tuberculosis subclinical proportion (the proportion of prevalent tuberculosis cases that are asymptomatic) and tuberculosis prevalence ratio by socio economic status. For most targets, estimates from the 2020 WHO Global TB Report were used (World Health Organization, 2020).

In countries including the HIV structure, we had an additional 3 input parameters, related to HIV incidence and ART initiation, and an additional four targets: HIV prevalence, ART coverage, and tuberculosis incidence and mortality in people living with HIV.

The model running time was 10 seconds on average, with runs usually taking longer in countries including the HIV structure usually, compared to countries without the HIV structure.

For each target, a lower and an upper bound were specified. Parameter sets were considered to match the targets if all the model outcomes were between the bounds. For more details on model structure, parameters and targets, see Supporting Materials, section A, and (Clark et al., 2022).

Model calibration was attempted for 92 countries without HIV structure and 23 countries including HIV structure, where all the data required to calibrate the model were available.

## 3. Calibration Method

In this section, we briefly describe how history matching with emulation works. We then show in detail how such a method was implemented in the case of our model.

### History matching with emulation

History matching concerns the problem of exploring the input space (i.e. all possible parameter sets) and identifying parameter sets that may give rise to acceptable matches between the model output and the empirical data. This part of the input space is referred to as *non-implausible*, while its complement is referred to as *implausible*. History matching proceeds as a series of iterations, called *waves*, where implausible areas of input space are identified and discarded. To identify those areas, we use emulators: statistical approximations of the model outputs which are built using a manageable number of model evaluations, and are typically several orders of magnitude faster than the model. If *D* is a set of model runs, we can use *D* to build an emulator for model output *f*. The emulator then provides us with a prediction *E*_*D*_[*f*(*x*)] for the value of *f* at parameter set *x*, and quantifies the uncertainty associated with that prediction, *Var*_*D*_[*f*(*x*)] (Andrianakis et al., 2017; Vernon et al., 2010). This information is then used to calculate the implausibility measure at the parameter set *x* for the model output *f*:

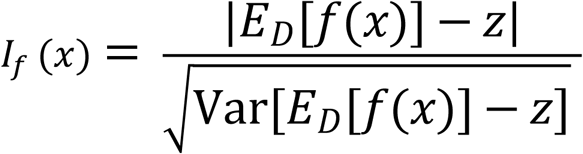

where *z* is the target for the model output *f*, and the variance in the denominator includes the uncertainty *Var*_*D*_[*f*(*x*)] in the emulator prediction and any other relevant form of uncertainty, e.g. the observation uncertainty or any structural discrepancy (accounting for the fact that no model perfectly represents reality). A high value of *I*_*f*_ (*x*) suggests that, even while factoring in all sources of uncertainty, the emulator prediction *E*_*D*_[*f*(*x*)] and the target *z* are too distant for *f*(*x*) to plausibly match the empirical data (Vernon et al., 2010).

For more details about emulators and the implausibility measure see Supporting Materials C. Figure 1 shows the various steps of the process.

**Fig 1:**
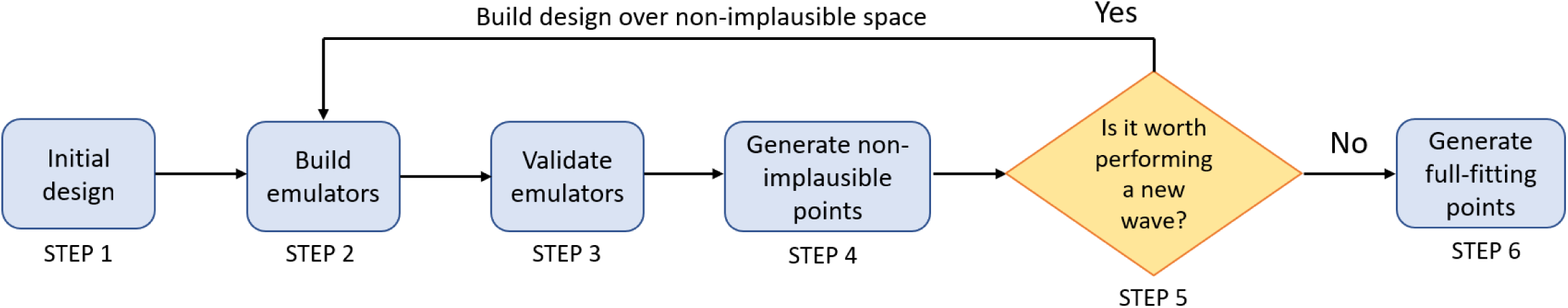
Workflow of history matching with emulation.

**Step 1**: We first run the model on the initial design points, a manageable number of parameter sets that are spread out uniformly across the input space, in accordance with established design principles (Vernon et al., 2010).

**Step 2**: The obtained model outputs provide us with training data to build emulators. The *hmer* package follows a Bayes Linear approach (Goldstein, 2012) for the construction of emulators.

**Step 3**: We test the emulators to check whether they are good approximations of the model outputs. Emulators which do not perform well enough are either modified and made more conservative, or omitted from the current wave.

**Step 4**: We generate a set of non-implausible parameter sets. To do this, we evaluate the emulators on a large number of parameter sets and we discard the implausible ones, i.e., those with implausibility measure above a chosen threshold.

**Step 5**: As this is an iterative procedure, each time we complete Step 4 we need to decide whether to perform a new wave or stop. In the former case, the model is run on the non-implausible parameter sets found in Step 4 and the process returns to Step 2. In the latter case, we progress to Step 6. One possible stopping criterion is if the uncertainty of the emulators is smaller than the uncertainty in the targets, indicating that further improving the performance of the emulators will make little difference to the rate of points generation. We may also choose to stop the iterations when we get emulators that provide us with points matching all targets at a sufficiently high rate. If our aim is to do forecasting, we would usually be happy to stop if we have found good matches and all of the areas of the non-implausible space produce roughly similar forecasts for key future outcomes of interest. Conversely, if there is substantial variation for key forecasts in the non-implausible space, then we would want to continue, even if we have found plenty of matches already. Finally, we might end up with all the input space deemed implausible at the end of a wave: this would raise doubts about the adequacy of the chosen model, or input and/or output ranges.

**Step 6:** We use the emulators to generate non-implausible points, we run the model on them and we retain only those points that match all targets.

For a more detailed introduction to history matching with emulation we refer the reader to Andrianakis et al., 2015; Vernon et al., 2010, 2018. For a direct comparison with ABC see McKinley et al., 2018.

### Performing history matching with emulation on the tuberculosis model

The *hmer* package, released on CRAN on April 14th 2022, contains a range of options at each step of the process, allowing the calibration process to be customised at each wave. As we were calibrating the model to a large number of countries, we used an automated approach, which did not require any customisation choices across different countries or waves. The calibration of each country was set to run for at most a week, with the aim of finding as many full fitting points as possible. We describe below how each step of the process was implemented.

**Step 1:** The model was first run at 400 input points, which were chosen through Latin Hypercube sampling. Of these, half were used to build the emulators (training points) and half to validate them (validation points).

**Step 2:** The model outputs at training points were then used to train the emulators.

**Step 3:** The model outputs at validation points were then used by the *hmer* package to conduct two diagnostic tests on emulators. The first assessed how accurately emulators reflected model outputs.

In Fig. 2, the emulator expectation (*E*_*D*_[*f*(*x*)]) is plotted against the model output (*f*(*x*)) for each validation point, providing the dots in the graph, with 95% credible intervals. Points for which the model output is outside the interval are in red. The exception to this is in places where the model output is far away from the target we want to match, as it is not important that the emulators are accurate in such places. An ‘ideal’ emulator would exactly reproduce the model results: this behaviour is represented by the green line. This diagnostic was used as an initial filter, to identify emulators that performed very poorly: if more than 25% of all validation points were in red, the emulator was discarded, and the corresponding output was not used to rule out implausible points for that wave. In the example shown above, the test was clearly successful (only one validation point in red), and the emulator proceeded to the second diagnostic.

**Fig. 2:**
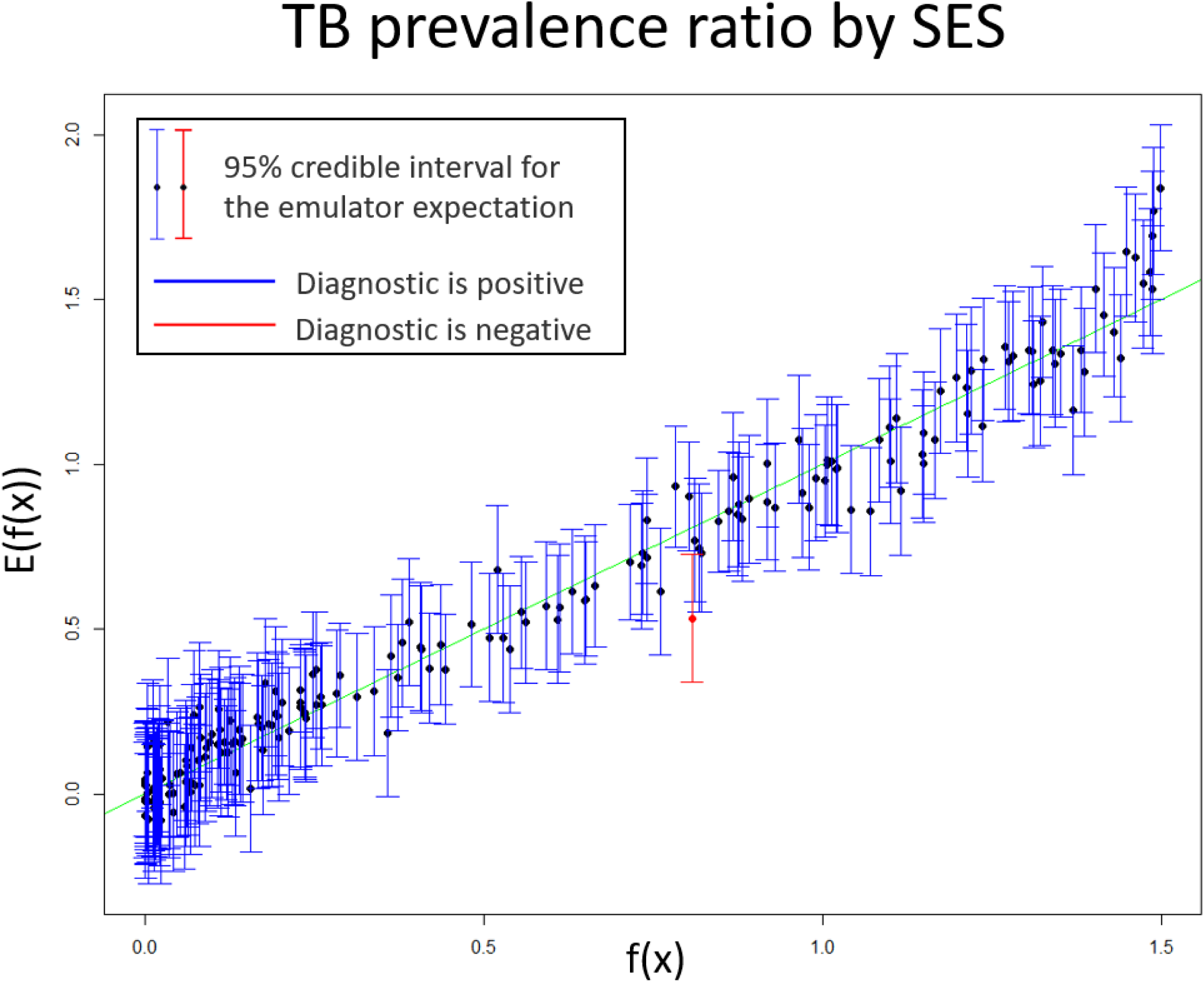
An example of the first emulator diagnostic. The emulator expectation (E[f(x)]) is plotted against the model output (f(x)) for each validation point, providing the dots in the graph. A 95% credible interval for the emulator expectation at each validation point is shown in the form of a vertical interval. Points for which the model output is outside the interval provided by the emulator are highlighted in red. The lower the percentage of points in red, the more accurate the emulator’s approximation of the model output.

Fig. 3 compares the emulator implausibility to the equivalent simulator implausibility (i.e. the implausibility calculated by replacing the emulator output with the model output). There are three cases to consider:

- The emulator and model both classify a set as implausible (bottom-left quadrant) or non-implausible (top-right quadrant). This is fine. Both are giving the same classification for the parameter set.
- The emulator classifies a set as non-implausible, while the model rules it out (top-left quadrant). This is also fine. The emulator should not be expected to shrink the input space as much as the simulator does, at least not on a single wave. Parameter sets classified in this way will survive this wave, but may be removed on subsequent waves as the emulators grow more accurate on a reduced input space.
- The emulator rules out a set, but the model does not (bottom-right quadrant): these points are highlighted in red and are the problematic sets, suggesting that the emulator is ruling out parts of the input space that it should not be ruling out.

**Fig. 3:**
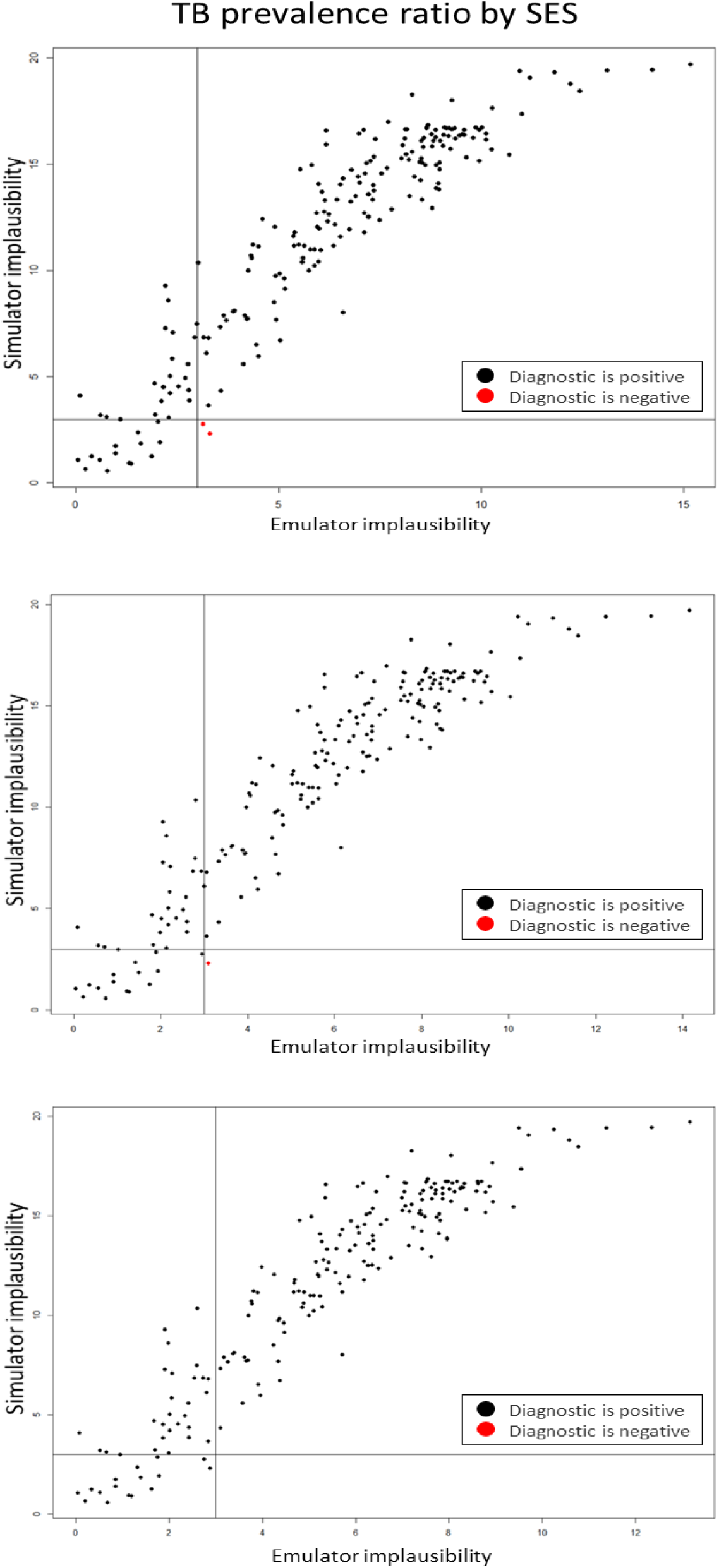
An example of the second emulator diagnostic. The simulator implausibility is plotted against the emulator implausibility. The top plot refers to the emulator for the TB prevalence ratio by SES as first trained. The middle plot shows the diagnostic after the emulator uncertainty was increased by 10%. The bottom plot shows the diagnostic after a further 10% increase of the emulator uncertainty

To ensure that no part of the input space was unduly ruled out, we required that no point was in red. We achieved this by iteratively increasing the emulator uncertainty by 10% (i.e. the length of the blue vertical bars in the first diagnostic), resulting in an emulator that was more cautious when ruling points out. In Fig. 3, increasing the uncertainty by 10% once removed one of the two red points (middle plot), and a further increase resulted in no red points (bottom plot).

**Step 4:** A new set of 400 non-implausible points was generated. Using the *hmer* package, new points were generated according to the following strategy. An initial set of points was generated using a Latin Hypercube Design, rejecting implausible parameter sets (Fig. 4, top-left). Pairs of non-implausible parameter sets were then selected at random and more sets were sampled from lines going through them, to identify those that are close to the boundary (Fig. 4. top-right). Finally, using all non-implausible points and boundary points found so far as seeding points, more parameter sets were generated using importance sampling to attempt to fully cover the non-implausible region (Fig. 4 bottom-left).

**Fig. 4:**
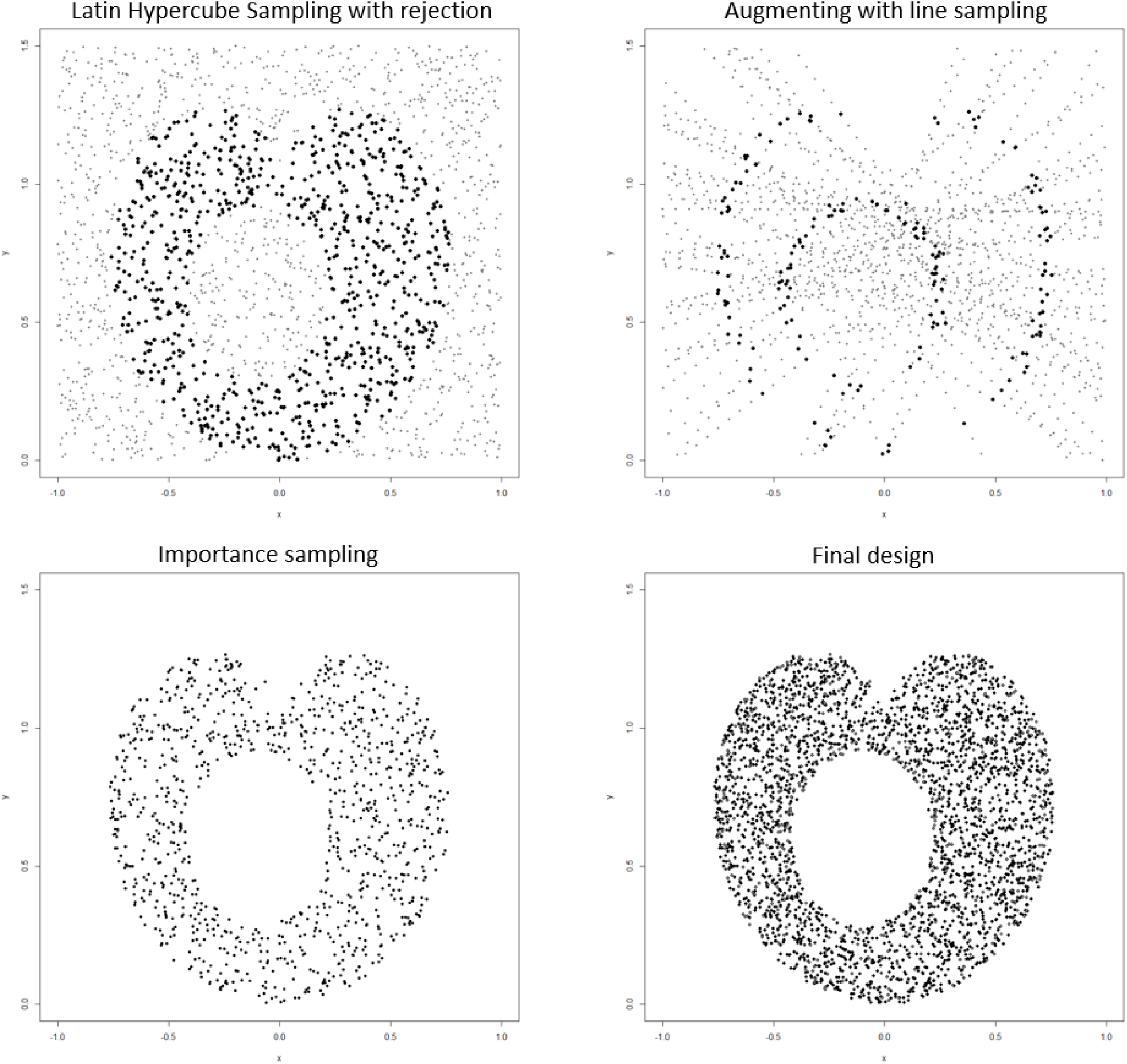
Strategy used to generate non-implausible parameter sets. The top-left graph shows an example of Latin Hypercube sampling, with non-implausible points in black and implausible points in grey. Line sampling was then performed (top-right right): non-implausible points found by Latin Hypercube (in grey) were connected by straight lines and points on those lines were explored. Parameter sets proposed by line sampling are in grey if implausible, or if in the interior of the non-implausible region and in black if close to the boundary. Finally, importance sampling, using mixture distributions, was carried out (bottom-left graph) using the non-implausible points found through Latin Hypercube and Line Sampling as seeding points. The final design is shown on the bottom-right graph.

**Step 5:** If the emulator uncertainty was larger than the target uncertainty, another wave was performed: the 400 points generated in STEP 4 were split in training and validation sets, the model was run on them and the process returned to STEP 2. Once the uncertainty of emulators was smaller than the uncertainty in the targets, the process continued to STEP 6.

**Step 6:** Rejection sampling was performed at this stage: a set of points was uniformly sampled from the non-implausible region using all available emulators. The model was then run on such points, and those matching all targets were accepted. We repeated this process, stopping when seven full days had passed since the model calibration process started.

### Derivative emulation

The calibration process described above could fail to find fully fitting points for one of two reasons: because no such points existed, or because history matching had not found them. Working with tools from the *hmer* package, we analysed countries where the fitting process failed, to identify the reason(s) why calibration could not be completed. We describe one of these tools, derivative emulation, here.

The term derivative emulation refers to the process of estimating the derivative of model outputs using the corresponding emulators. More precisely, once an emulator is trained for a model output *f*, we can use it to get a prediction 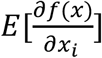 of the partial derivative of *f* with respect to the i-th parameter at any given parameter set *x*, with associated uncertainty 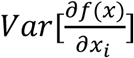. This is a low cost and potentially powerful approach, since it does not require any additional model evaluations. The hmer package has a function that, given a set of emulators and a parameter set *x* not fitting all targets, estimates the partial derivatives of all model outputs at *x*. It then uses the gradient descent method to find the best direction to move along in order to improve (or at least not worsen) all emulator evaluations simultaneously, and then proposes a new parameter set. This function allows us to explore parameter sets near *x*, to determine if any of those are a better fit to the data.

## 4. Results

Of the 115 countries that we analysed, 105 were fully calibrated to all targets. In the time available, the number of parameter sets hitting all targets found by history matching with emulation varied by country, from a minimum of 232 to a maximum of 8857. For 101 of the 105 calibrated countries, at least 1000 full-fitting points were found.

We now show some visualisations produced using the hmer package for one of the calibrated countries. Fig. 5 shows (log-scaled) output values for non-implausible parameter sets at each wave. The plot clearly shows how the performance of non-implausible parameter sets improves each wave, with an increasing proportion of output values falling with the target ranges. In Fig. 6, output values for non-implausible parameter sets at each wave are shown for each combination of two outputs. For wave 7, we show only the fully fitting points. The main diagonal shows the distribution of each output at the end of each wave, with the vertical red lines indicating the lower and upper bounds of the target. Above and below the main diagonal are plots for each pair of targets, with rectangles indicating the target area where full fitting points should lie (the ranges are normalised in the figures above the diagonals). These graphs can provide additional information on output distributions, such as correlations between them.

**Fig. 5:**
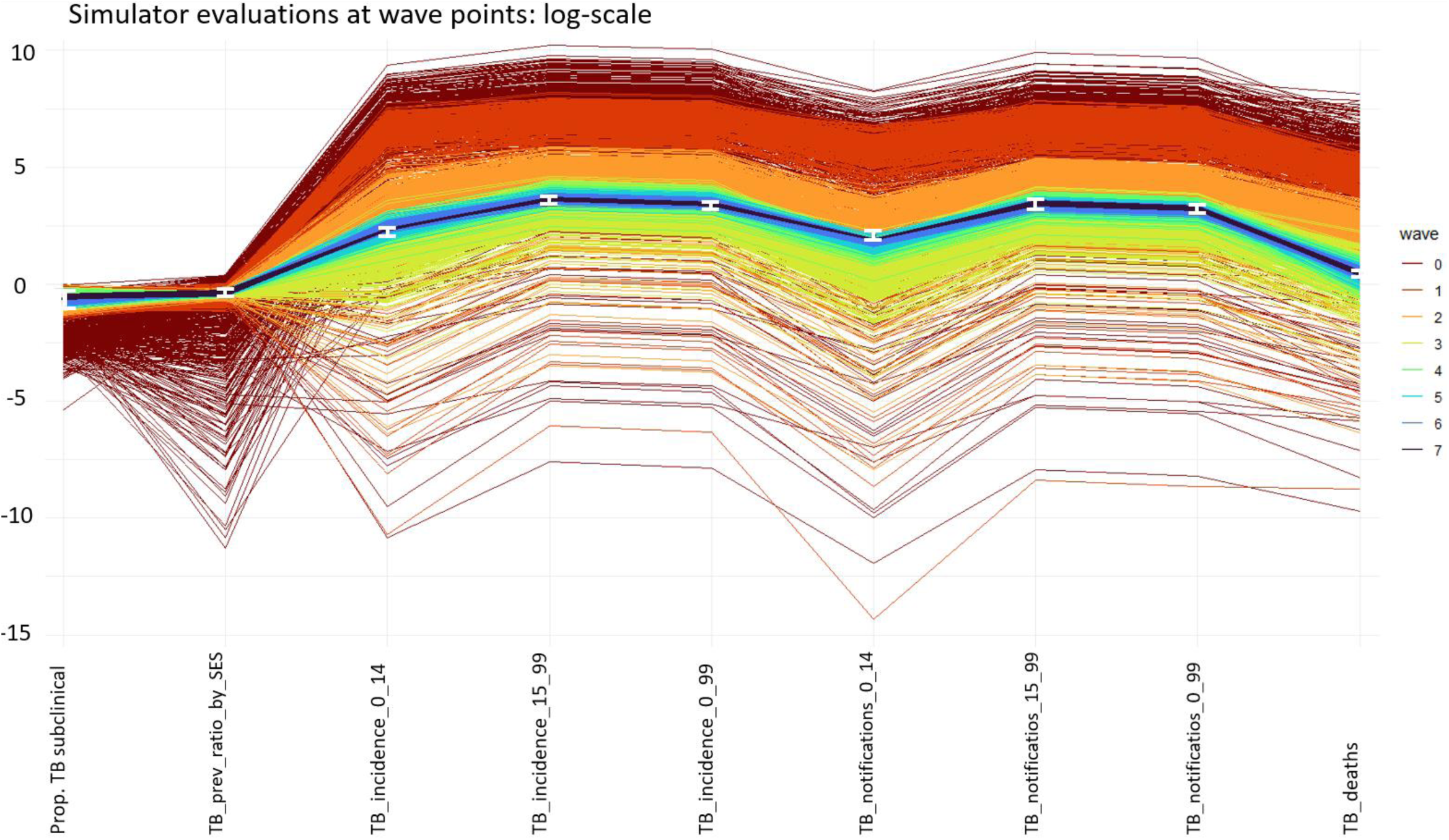
Plot of simulator evaluations at non-implausible parameter sets for one of the calibrated countries. Each column refers to a model output, and parameter sets are coloured based on the wave they belong to, with darker colours for later waves. The model outputs (y-axis) were log-scaled to improve the readability of the graph. For each model output the lower and upper bounds of the correspondent target are shown as the white error bars.

**Fig. 6:**
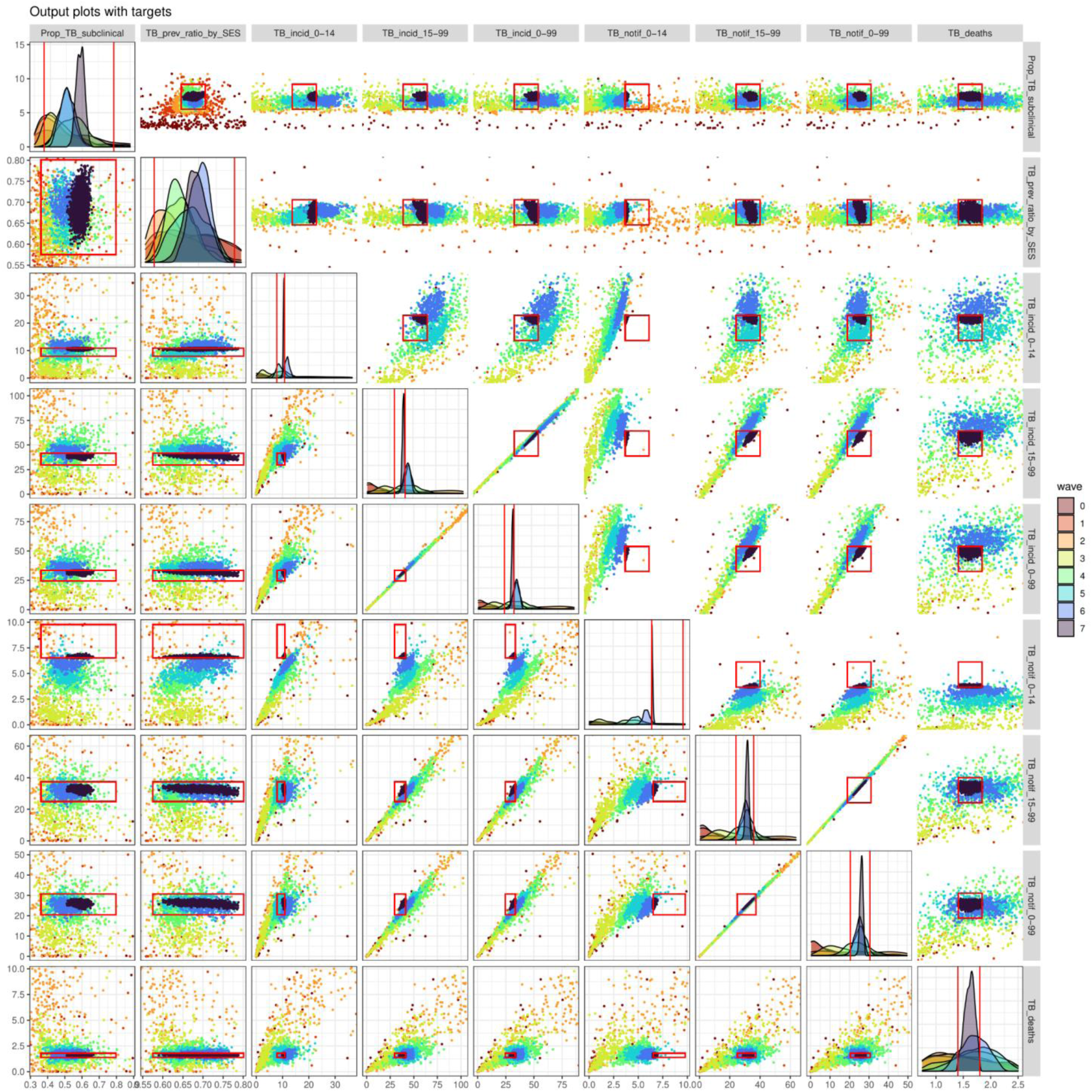
Pair plots of the outputs for non-implausible parameter sets in different waves, for the same country shown in Fig. 5 and Fig. 6. The main diagonal shows the distribution of each output, with the vertical red lines indicating the bounds of the target. Below the main diagonal are plots for each pair of targets, with rectangles indicating the target area where full fitting points should lie. Above the diagonal we have similar plots, but the ranges have been normalised, so that the target area where full fitting points should lie is now a red square centred at the middle of the plot.

The third visualisation (Fig. 7) shows the distribution of the non-implausible input parameter space each wave. The plots in the main diagonal show the distribution of each parameter, which tends to narrow each wave. The off-diagonal boxes show plots for all possible pairs of parameters. These visualisations give us information about the extent to which each of the parameters has been constrained, and about correlations between parameters. For example in Fig. 7 the probability of transmission and the relapse rate have an extremely narrow posterior, while the background mortality rate has a much wider posterior. The plot also shows the presence of a negative correlation between the TB disease treatment rate and the *Mtb* infection self-clearance rate. Overall, for each country, history matching with emulation greatly reduced the input space to be searched for full-fitting points, with an estimated median reduction factor of 6 × 10^8^ (see supporting materials, section D for more details). Fig 8 shows a histogram of the log10-transformed reduction factors for all countries.

**Fig. 7:**
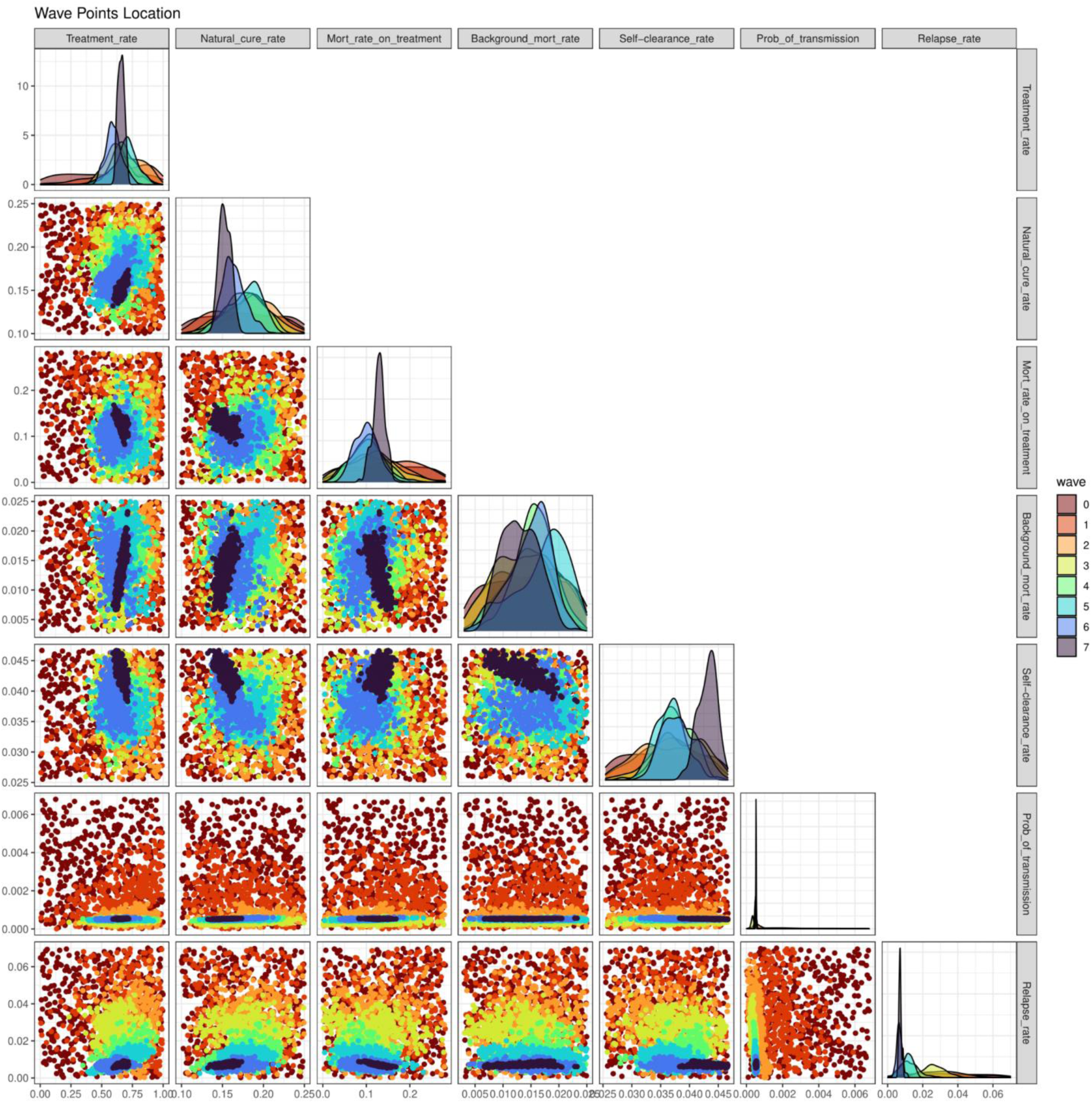
Pair plots of the non-implausible parameter sets wave after wave, for the same country shown in Fig. 5. The plots in the main diagonal show the distribution of each parameter, while in the off-diagonal boxes we have plots for all possible pairs of parameters. Each colour corresponds to a specific wave, with darker colours representing later waves.

**Fig 8:**
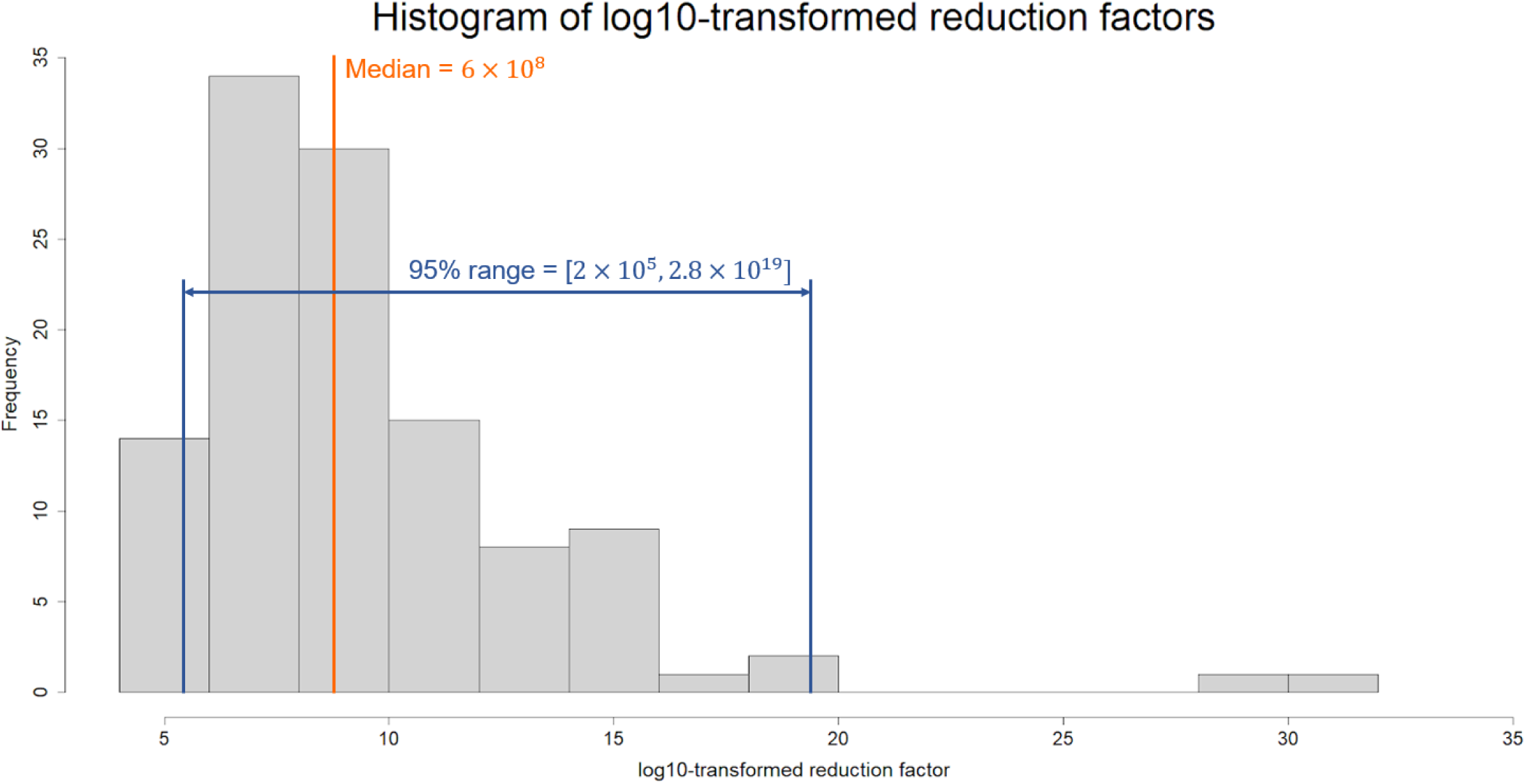
For each of the 115 countries we compared the initial input space and the final non-implausible space, estimating the correspondent reduction factor. The plot shows a histogram of the log10-transformed reduction factors for all countries. The estimated median reduction factor was 6 × 10^8^, with 95% of all reduction factors lying in the interval [2 × 10^5^, 2.8 × 10^19^].

### Computational cost of the calibration process

Each country was set to run on a single computer node (typically a 64bit Intel Xeon processor at 2.6 GHz) for 7 full days, with the aim of finding as many full-fitting points as possible. For many countries, hundreds of full-fitting points were identified well before the end of the 7 allocated days. On average across all countries, model running time accounted for 25% of the total computational cost, emulator training/diagnostic time for 5%, and points proposal time for 70%. (Note that this study was conducted using a developmental version of the package and point generation is now around 5 times faster in Version 1.0.0 of the package. Future improvements are planned, to further optimise the generation of non-implausible points.) The points proposal time increased linearly with waves, as all previous emulators needed to be evaluated each wave. On average, 5,000–10,000 points were investigated at each wave to find 400 non-implausible ones. Note that such a huge amount of evaluations were only possible thanks to the high speed of emulators: while a model run took 10 seconds on average, evaluating an emulator took just a few milliseconds.

### Analysis of countries that could not be fitted to data

In 9 uncalibrated countries (7 countries without HIV structure and 2 countries with HIV structure), we used *hmer* package visualisation tools to understand why the calibration process could not be completed. Let us consider country A, for which history matching found points matching 7 out of 9 targets at most. Figure 9 (left plot) shows tuberculosis notifications in adults (ages 15–99) in 2019 and tuberculosis notifications in young individuals (ages 0–14) in 2019, with the red rectangle indicating the target area.

**Fig 9:**
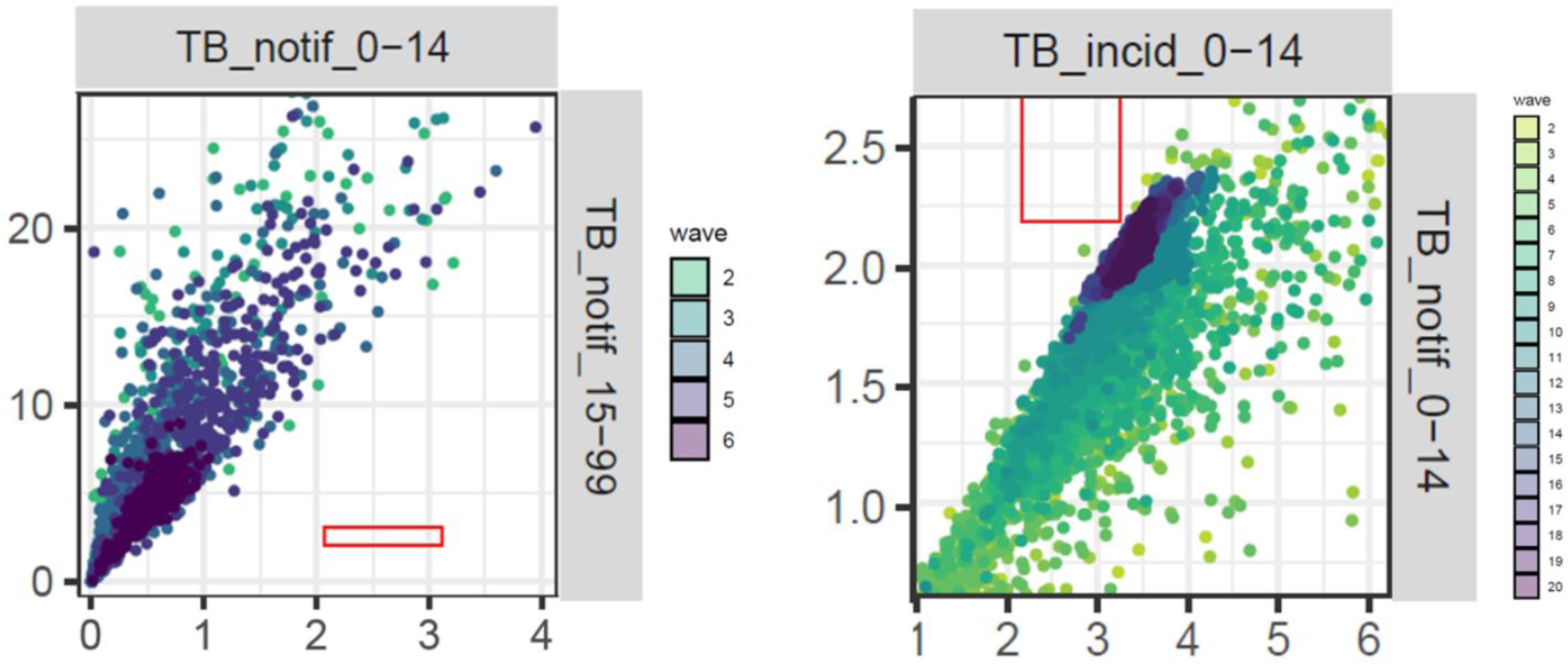
On the left, pair plot of TB notifications in young individuals against TB notifications in adults, for parameter sets in waves 2, 3, 4, 5, 6 (country A). On the right, pair plot of TB incidence in young individuals and TB deaths for parameter sets in waves 2, 3,…, 19 (country F).

We see that the two outputs are highly correlated and that, in all waves, all points lie far from the red rectangle. This suggests that our model is not able to match the two targets simultaneously. Four more countries showed a similar clear incompatibility between two (or more) targets (see supporting material section B).

The remaining four countries (referred to as countries F, G, H, I) that could not be fitted to data also showed some evidence of an incompatibility between certain pairs of targets, although in a weaker form. For example, in country F, our model seemed incapable of matching overall tuberculosis notifications and tuberculosis disease incidence in young individuals (Fig. 9, right plot, where we see a “wall” in output space that prevents the model outputs from reaching the red target square. Such a wall is likely due to inherent and possibly non-obvious constraints in the model). The bounds for these two targets were almost identical. As our model did not include false positive notifications, this meant that nearly all TB in children would need to be diagnosed and notified for the model to fit the targets, which was incompatible with the plausible input ranges and other output target ranges used in the calibration.

Similar plots for countries G, H and I can be found in Supporting Material, Section B. Since the incompatibility between outputs was present but less pronounced in countries F, G, H and I, one could wonder if by moving around the last-wave points - which were often close to matching the potentially incompatible outputs - one could find full fitting points. We explored this using derivative emulation.

For countries G, H and I, the maximum number of targets met by a point in the last wave was 8 (out of 9). For these countries, even though derivative emulation increased the number of targets that some parameter sets matched, it could not find any parameter sets hitting all 9 targets. For country F, neither last wave points nor points found by derivative emulation matched more than 7 (out of 9) targets.

Finally, one country had inconsistencies in the data that made calibration unachievable.

## 5. Discussion

In this paper, we have demonstrated how the *hmer* R package allowed us to perform history matching with emulation on a complex deterministic tuberculosis model to a total of 115 low-and middle-income countries. For each country, the model was fit to 9-13 target measures, by varying 19-22 input parameters. Overall, 105 countries were successfully calibrated, while for ten countries history matching with emulation could not identify any full-fitting parameter sets. Such countries were analysed further, through derivative emulation and visualisations tools offered by the *hmer* package.

Calibrating 105 countries in just a few days was possible thanks to *hmer*, which allowed us to implement history matching with emulation in a straightforward and effective way. Using this method, we were able to fully explore the input space, and to identify a set of points that represent all possible conditions under which our model could match the data. In nine countries that could not be fitted to data, visualisation tools, paired with derivative emulation methods where necessary, provided us with evidence that calibration of the model was unlikely to be possible with the given input and output ranges.

Unlike most other full calibration methods, which attempt to make probabilistic statements about the posterior distribution over the input space, history matching can work with expectation and variances only, and aims to discard implausible areas of the input space. This, and the use of emulators, mean that the calculations involved in history matching tend to be more tractable and straightforward to implement. Furthermore, since the process is iterative, it is not necessary to work with all inputs and outputs simultaneously: for example, if an output cannot be emulated accurately in initial waves, it can be put aside and emulated in later waves, as emulation usually becomes easier as the waves progress (Vernon et al., 2010). All these characteristics make history matching a useful method for calibrating complex models. In addition to being a useful calibration method in its own right, when we are interested in making probabilistic statements about the posterior of the model’s parameters, history matching can be used as a pre-calibration procedure, in conjunction with probabilistic calibration techniques (Vernon et al., 2018, Vernon et al., 2010). While methods such as Approximate Bayesian Computation or Bayesian model calibration may struggle to adequately explore the large input space of a multi-output model, their application on the greatly reduced space that is the output of history matching can produce hybrid methods that are more tractable and combine the strengths of both approaches.

Multi-output models, such as the one in this article, very often exhibit correlation between their outputs. This aspect was partially incorporated by building accurate 1-dimensional emulators that together captured the joint behaviour of the model outputs. However, full multivariate emulation would bring further benefits. Taking output correlation fully into account would improve the emulation process and subsequently the performance of history matching, but requires more sophisticated emulators, as well as a more detailed structure for model and observation uncertainty which takes into account these correlations. These features, together with variance emulation, were not available at the time of this work, but are now part of the standard functionalities of version 1.0.0 of the *hmer* package. In particular, these features make it possible to use *hmer* to calibrate stochastic models, without having to average over multiple model runs for each parameter set. Also note that point generation is around 5 times faster in version 1.0.0 than it was in the developmental version of the package that was used in this work. Future improvements are planned, to further optimise the generation of non-implausible points. Features to be included in subsequent versions of the package will allow the user to address data quality problems, compare different models, and use emulators to make forecasts.

In conclusion, this work shows that the *hmer* package can play a key role in making history matching with emulation accessible to the community of epidemiologists, facilitating fast and efficient model calibration. By addressing the current lack of methodologies to robustly calibrate complex models and perform uncertainty analysis on them, *hmer* constitutes an invaluable addition to the epidemiologist’s calibration tool-kit.

## Data Availability

All data produced in the present study are available upon reasonable request to the authors.

## Supporting Materials

### A. Description of the calibration task

We describe the parameters varying in the calibration process and the set of targets we fitted our model to. In order to do so, we need to briefly describe the compartments of our model. For full details see Clark et al., 2022.

#### TB Natural History Dimension

In our model, the population was assigned to the compartments described in Table A1 below based on their TB status. The TB natural history model is specified in Figure A1 below.

**Table A1.** Compartments included in the TB natural history structure

| Symbol | Compartment Name | Compartment Description |
| --- | --- | --- |
| $U_N$ | **Uninfected - Naive** | Individuals with no previous exposure to *Mtb* |
| $U_C$ | **Uninfected - Cleared** | Individuals *with* previous exposure/infection who were able to self-clear. No viable bacteria remaining. |
| $I_F$ | **Infection - Fast** | Infected individuals who are at risk of either fast progressing to active disease within 2 years or continuing to the $I_S$ class |
| $I_S$ | **Infection - Slow** | Infected individuals who did not fast progress within 2 years, but remain at risk of progressing to active disease or self-clearing or reinfecting back to $I_F$ |
| $D_s$ | **Subclinical Disease** | Individuals with active, infectious TB disease, but no symptoms |
| $D_c$ | **Clinical Disease** | Individuals with active, infectious TB disease, with symptoms |
| $T$ | **On-treatment** | Individuals detected from $D_c$ and started on treatment. |
| $R$ | **Resolved** | Completed treatment or naturally cured from $D_s$ or $D_c$ |

**Figure A1.**
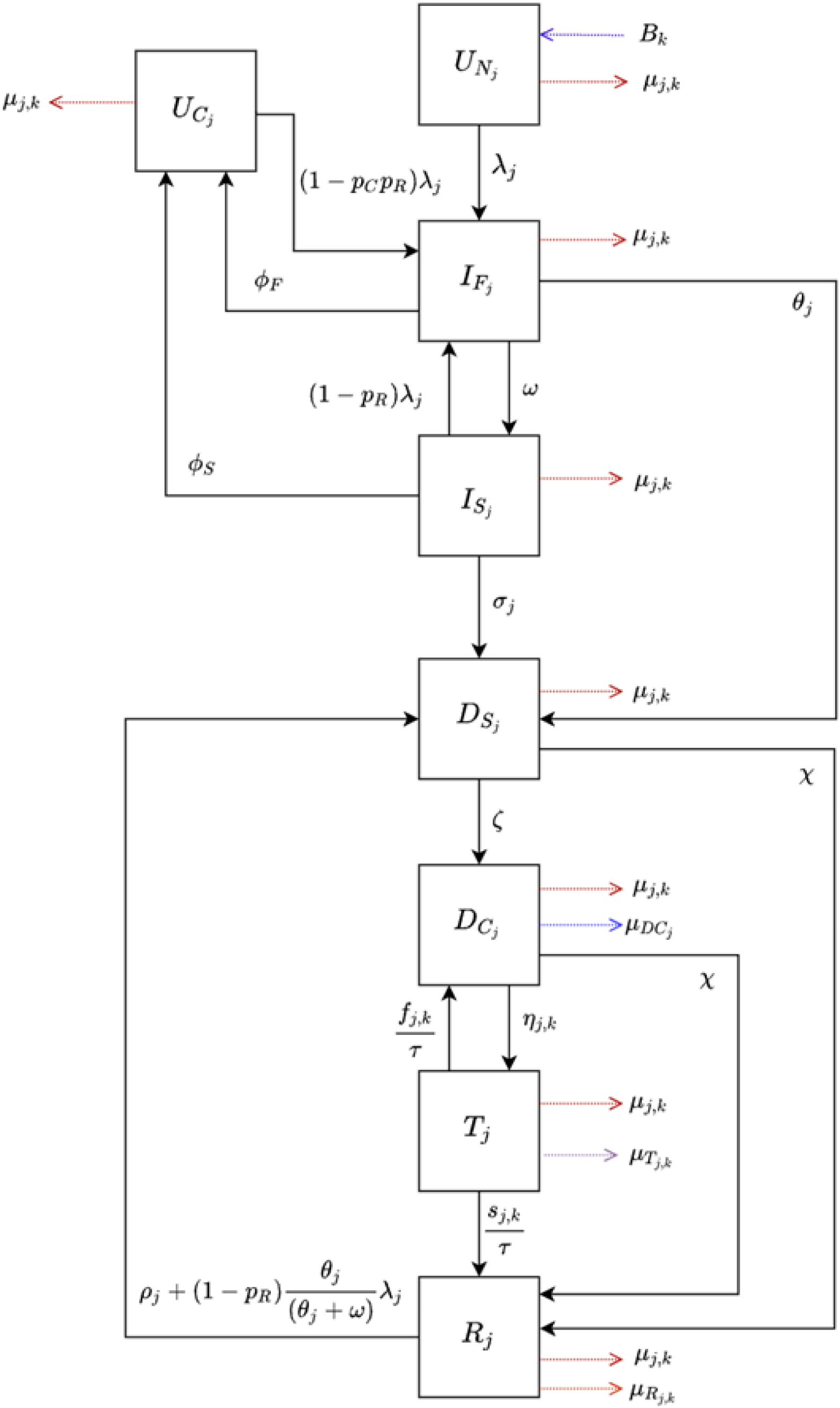
The TB natural history diagram

#### Input parameters and target outputs

For countries without HIV structure we had 19 varying parameters and 9 targets (all for the year 2019), as shown in the tables below.

**Table A2.** Description of parameters for countries without the HIV structure

| Parameter Description | Units | Symbol | Label on plots |
| --- | --- | --- | --- |
| **Natural history** |  |  |  |
| Rate of natural cure from Clinical Disease ($D_C$) and Subclinical Disease ($D_S$) | Per person, per year | $\chi$ | Natural cure rate |
| Rate of fast progression to disease | Per person, per year | $\theta$ | Fast progression rate |
| Age scaling parameter for the rate of fast progression ($\theta$) | - | $j_1$ | Age scaling theta |
| Rate of reactivation from Infection – Slow ($I_S$) | Per person, per year | $\sigma$ | Reactivation rate |
| Age scaling parameter for rate of reactivation from Infection – Slow ($\sigma$) | - | $j_2$ | Age scaling sigma |
| Rate of relapse from Resolved ($R$) | Per person, per year | $\rho$ | Relapse rate |
| Age scaling parameter for rate of relapse from Resolved ($\rho$) | - | $j_3$ | Age scaling rho |
| Rate of progression from Subclinical Disease ($D_S$) to Clinical Disease ($D_C$) | Per person, per year | $\zeta$ | Progression rate |
| Rate of self-clearance from Infection – Slow ($I_S$) to Uninfected – Cleared ($U_C$) | Per person, per year | $\phi_S$ | Self-clearance rate |
| Probability of transmission per infectious contact | - | $p_T$ | Prob. of transmission |
| Multiplier for TB prevalence | - | - | TB prev. multiplier |
| **Mortality** |  |  |  |
| Mortality rate from untreated clinical TB | Per person, per year | $\mu_{DC}$ | Mort. rate untreated |
| Background mortality rate | Per person, per year | $\mu_K$ | Background mort. rate |
| Mortality rate on treatment | Per person, per year | $k/\tau$ | Mort. rate on treatment |
| Age scaling parameter for mortality rates | - | $s_{Age}$ | Age scaling mort. rates |
| **Treatment** |  |  |  |
| Treatment duration | year | $\tau$ | Treatment duration |
| Rate of treatment initiation | Per person, per year | $\eta$ | Treatment rate |
| Age scaling parameter for rate of treatment initiation ($\eta$) | - | $j_4$ | Age scaling eta |
| **Protection** |  |  |  |
| High access-to-care* | - | $p_E$ | SES parameter |
| Protection from reinfection offered by Infection – Slow ($I_S$), Infection – Fast ($I_F$) and Resolved (R) | - | $p_R$ | Prot. against reinf. |

\*In the low access to care class, $p_E = 0$ . In the high access to care class, the progression rate from from Infection Fast and Infection slow to Subclinical Disease and the relapse rate from Recovered to Subclinical Disease are multiplied by $(1 - p_E)$ . The treatment initiation rate is instead divided by $(1 - p_E)$ in the high access to care class.

**Table A3.** Description of targets for countries without the HIV structure

| Target Description | Age group | Label on plots | In terms of the model |
| --- | --- | --- | --- |
| TB subclinical proportion | 0-99 | Prop_TB_subclinical | Ratio of $D_S$ to $(D_S + D_C)$ |
| TB prevalence rate ratio by access-to-care (SES) | 0-99 | TB_prev_ratio_by_SES | Ratio of $(D_S + D_C)_{High}$ to $(D_S + D_C)_{Low}$<br>High = high access to care strata<br>Low = the low access to care strata |
| TB incidence (per 100,000 population/year) | 0-14 | TB_incid_0-14 | All the in-flows into $D_S$, i.e., $(L_S \text{ to } D_S) + (L_S \text{ to } D_S) + (R \text{ to } D_S)$ |
| TB incidence (per 100,000 population/year) | 15-99 | TB_incid_15-99 |  |
| TB incidence (per 100,000 population/year) | 0-99 | TB_incid_0-99 |  |
| TB notifications (per 100,000 population/year) | 0-14 | TB_notif_0-14 | Flow from $D_C$ to T |
| TB notifications (per 100,000 population/year) | 15-99 | TB_notif_15-99 |  |
| TB notifications (per 100,000 population/year) | 0-99 | TB_notif_0-99 |  |
| TB mortality (per 100,000 population/year) | 0-99 | TB_deaths | All the in-flows into TBDead, i.e., $(D_C \text{ to } TBDead) + (T \text{ to } TBDead)$ |

Parameters for countries including the HIV structure include all parameters in Table A2 plus the following three in Table A4.

**Table A4.** Description of additional parameters specific to countries including the HIV structure

| Parameter description | Units | Symbol | Label on plots |
| --- | --- | --- | --- |
| Multiplier for ART incidence, varied to fit the 2019 HIV ART coverage | - | $\alpha_H$ | ART incid. multiplier |
| Multiplier for HIV incidence, varied to fit the 2019 HIV prevalence | - | $\lambda_H$ | HIV incid. multiplier |
| Multiplier for TB progression in HIV+, varied to fit the 2019 TB incidence rate in HIV positive population | - | $\theta_{mul}$ | TB-HIV progress. multiplier |

Targets for HIV countries include all targets in the non-HIV table plus the following four.

**Table A5.**
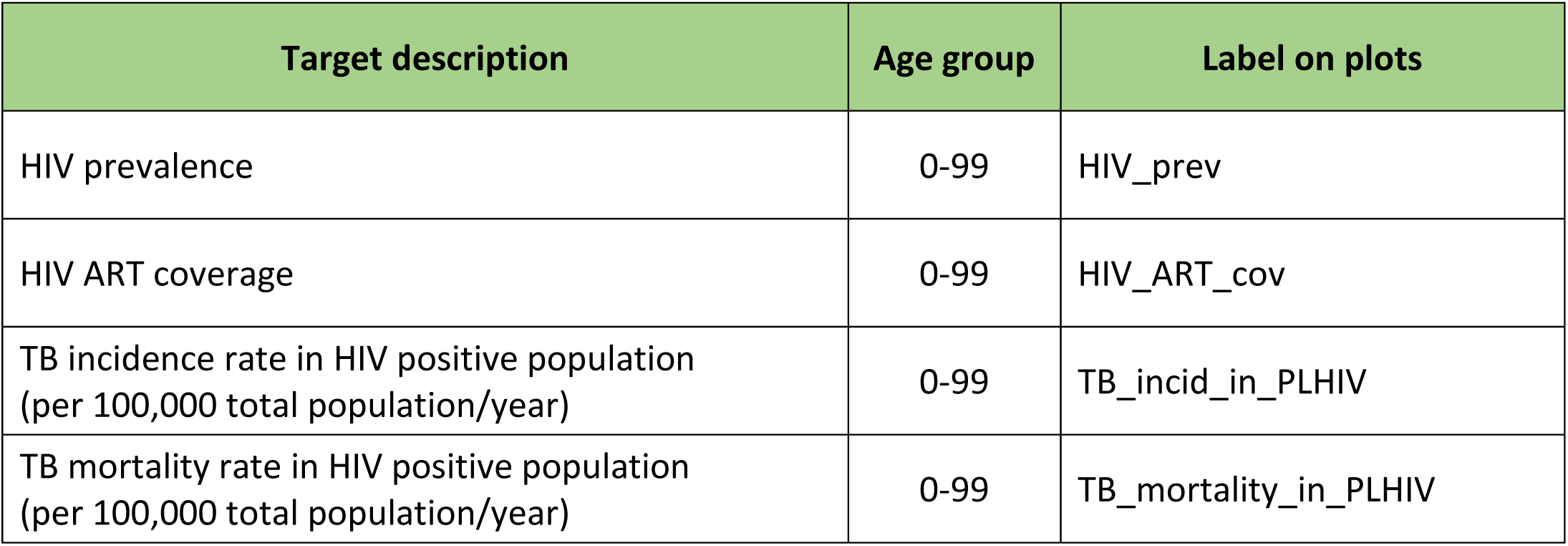
Description of targets specific to countries including the HIV structure

## B. Analysis of countries that could not be fitted to data

Apart from countries A and F, discussed in the main paper, seven more countries showed an incompatibility between two or more of the targets.

For countries B, C, D, E, we identified pairs of outputs that seemed strongly incompatible.

**Country B:**
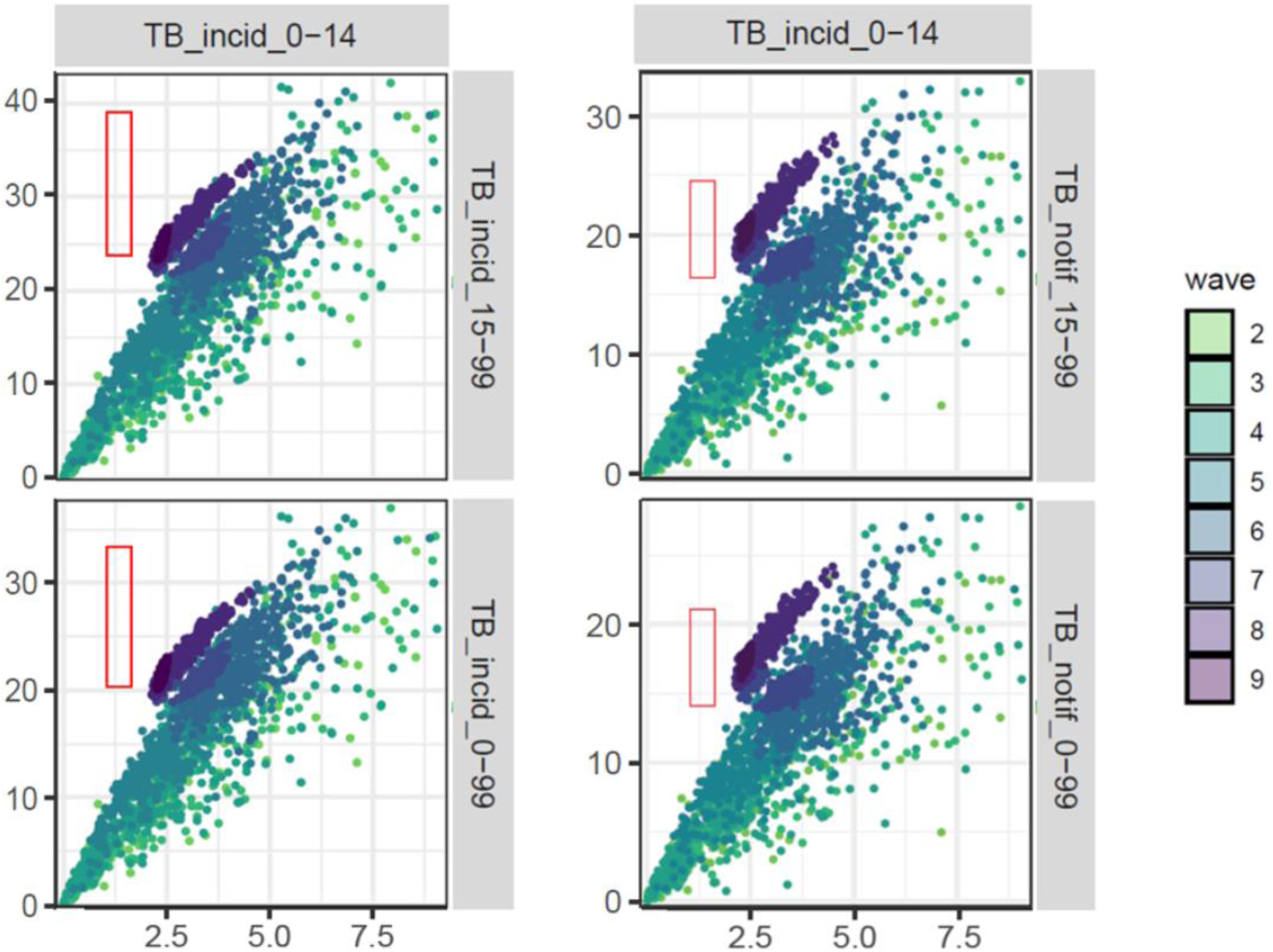
TB incidence in young people seem to be incompatible both with the other TB incidence targets (first column) and with TB notifications in adults and TB notifications in all the population (second column). Pair plot of TB incidence in young individuals against TB incidence and TB notifications in adults and in all the population, for parameter sets in waves 2, 3, 4, 5, 6, 7, 8, 9 (country B).

**Country C:**
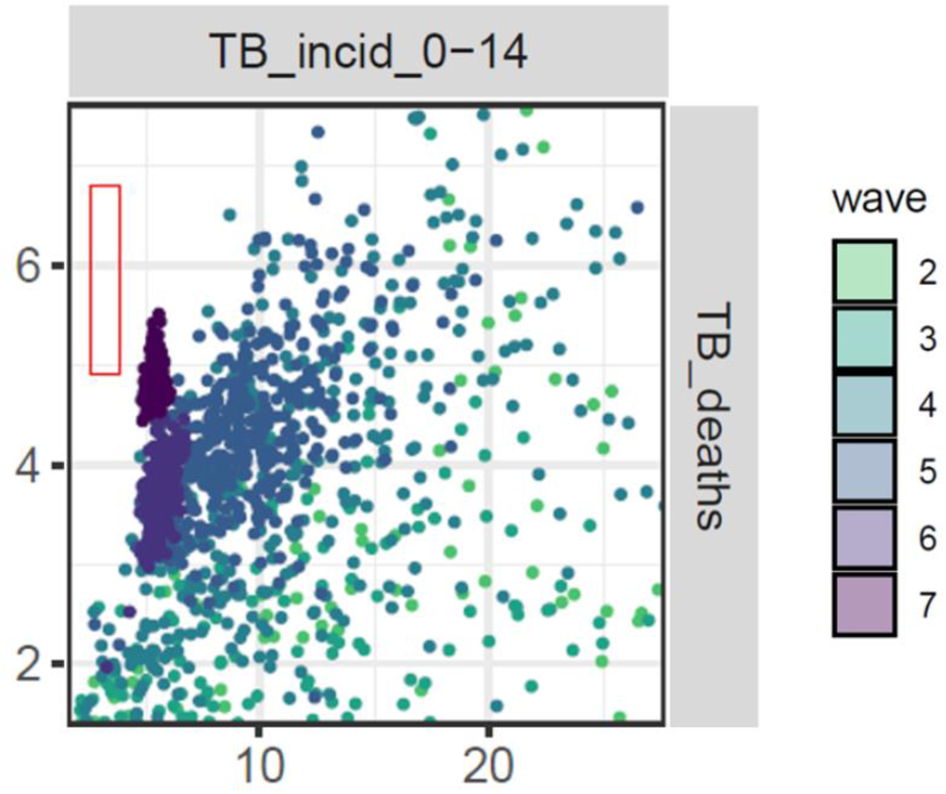
TB incidence in young people and TB deaths in all the population seem to be incompatible. Pair plot of TB incidence in young individuals against TB deaths in all the population, for parameter sets in waves 2, 3, 4, 5, 6, 7 (country C).

**Country D:**
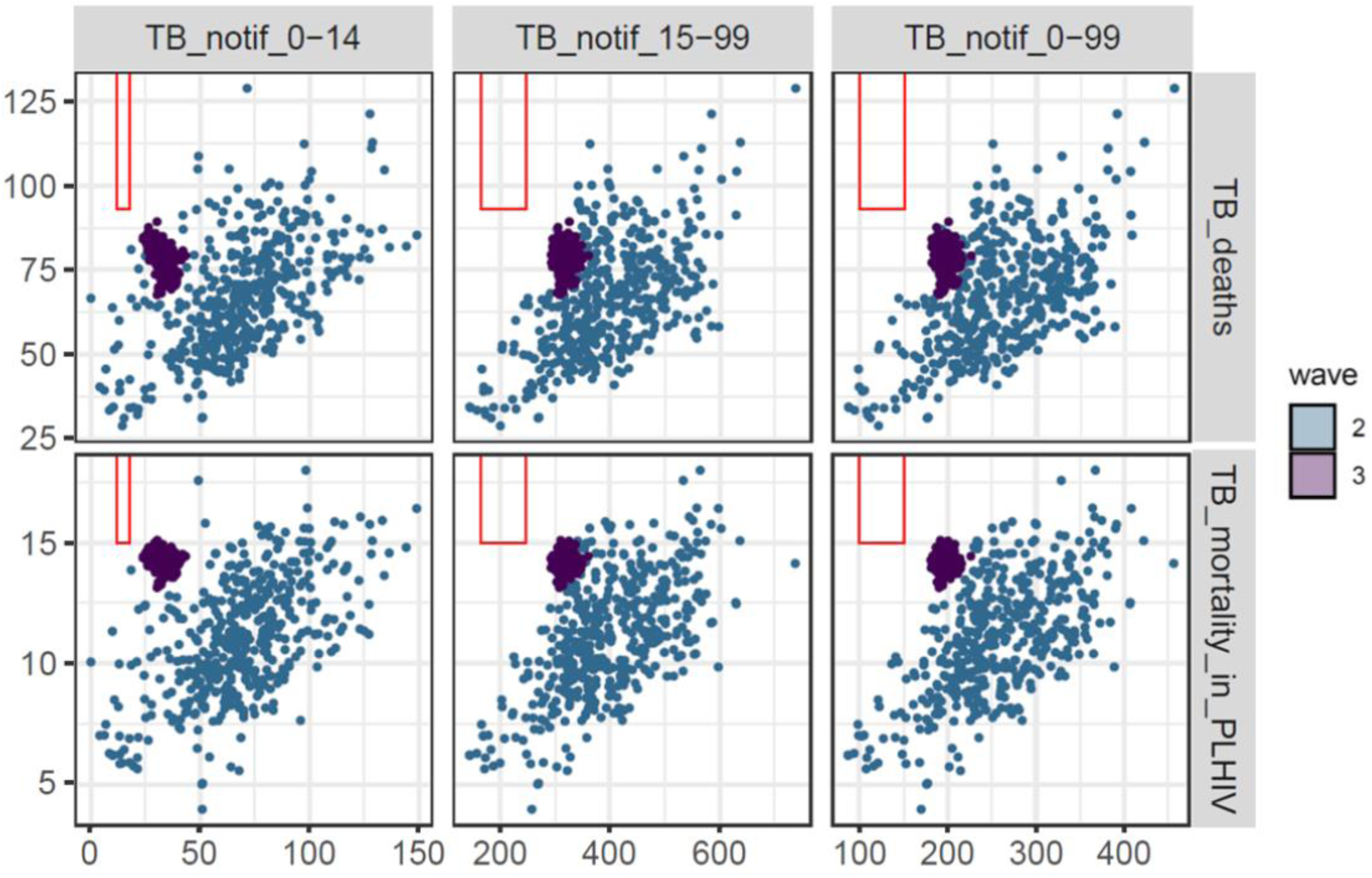
in this country with HIV structure, all three TB notifications targets seem to be incompatible with TB deaths and with TB mortality in people living with HIV. Pair plot of TB notifications in young individuals, adults and in all the population against TB deaths in all the population and TB mortality in PLHIV, for parameter sets in waves 2, 3 (country D).

**Country E:**
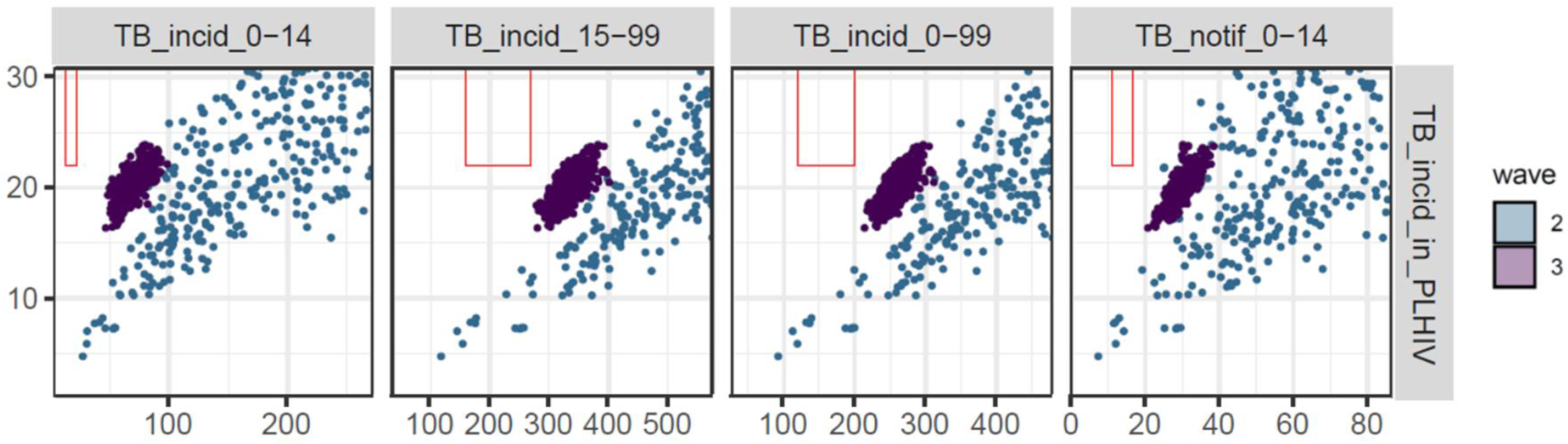
in this country with HIV structure, TB incidence in people living HIV seems to be incompatible with all three TB incidence targets and with TB notifications in young people. Pair plot of TB incidence in PLHIV against TB incidence (in young individuals, adults and in all the population) and TB notifications in young individuals, for parameter sets in waves 2, 3 (country E).

In countries G, H and I the incompatibility was less pronounced.

**Country G:**
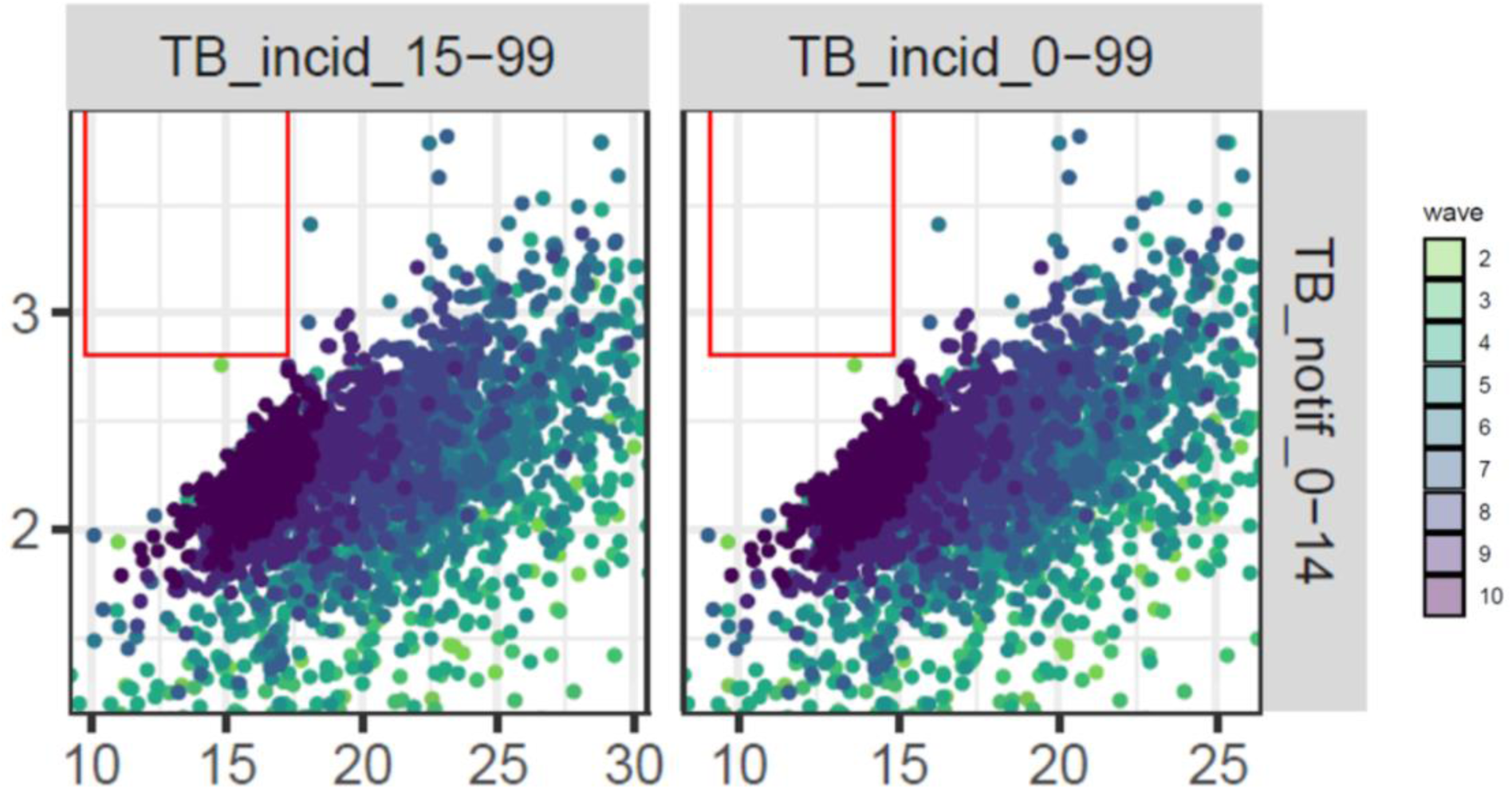
TB incidence in adults and in all the population are possibly incompatible with TB notifications in young individuals. Pair plot of TB incidence in adults and in all the population against TB notifications in young individuals, for parameter sets in waves 2, 3,…, 10 (country G).

**Country H:**
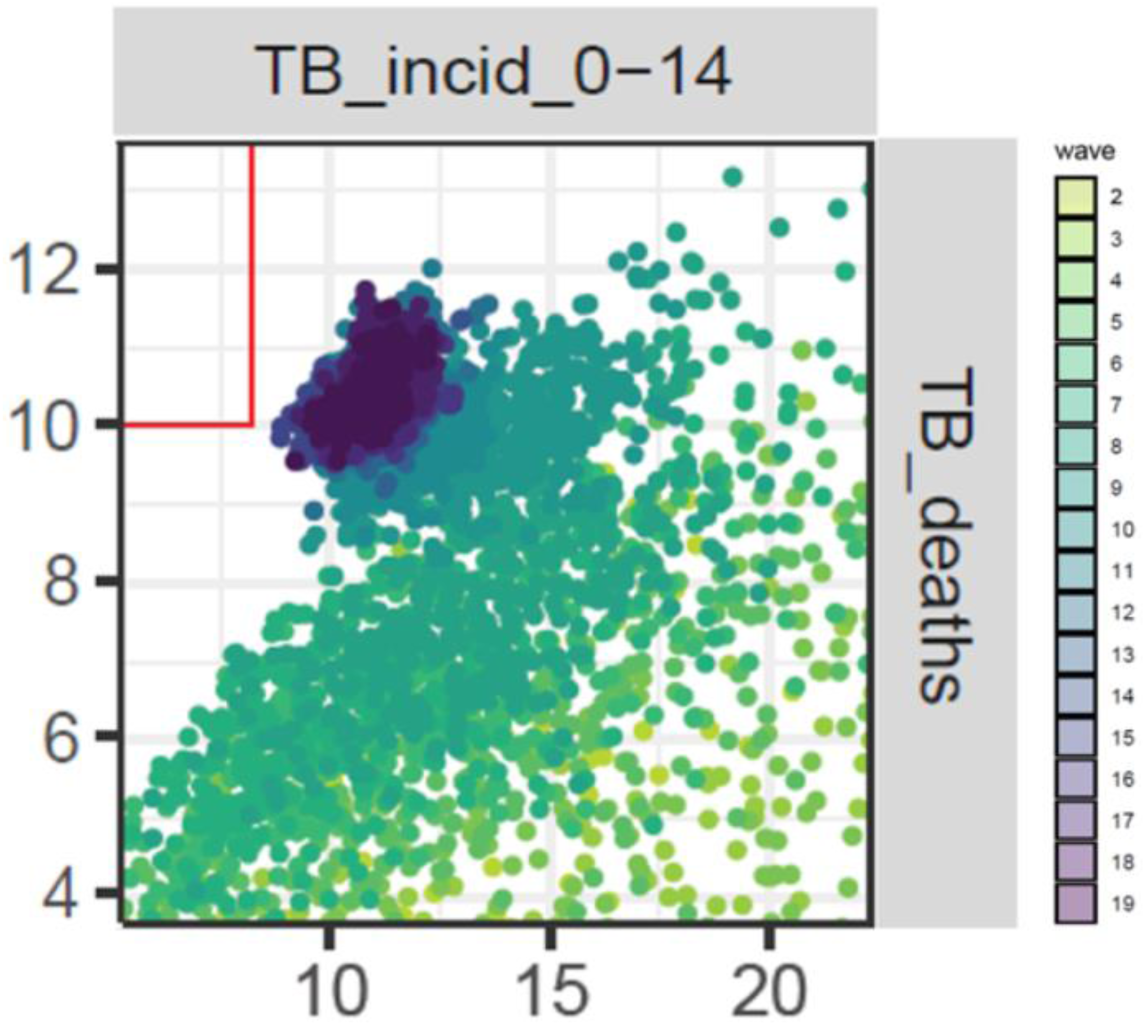
TB incidence in children is possibly incompatible with TB notifications in young individuals. Pair plot of TB incidence in young individuals and TB deaths for parameter sets in waves 2, 3,…, 19 (country H).

**Country I:**
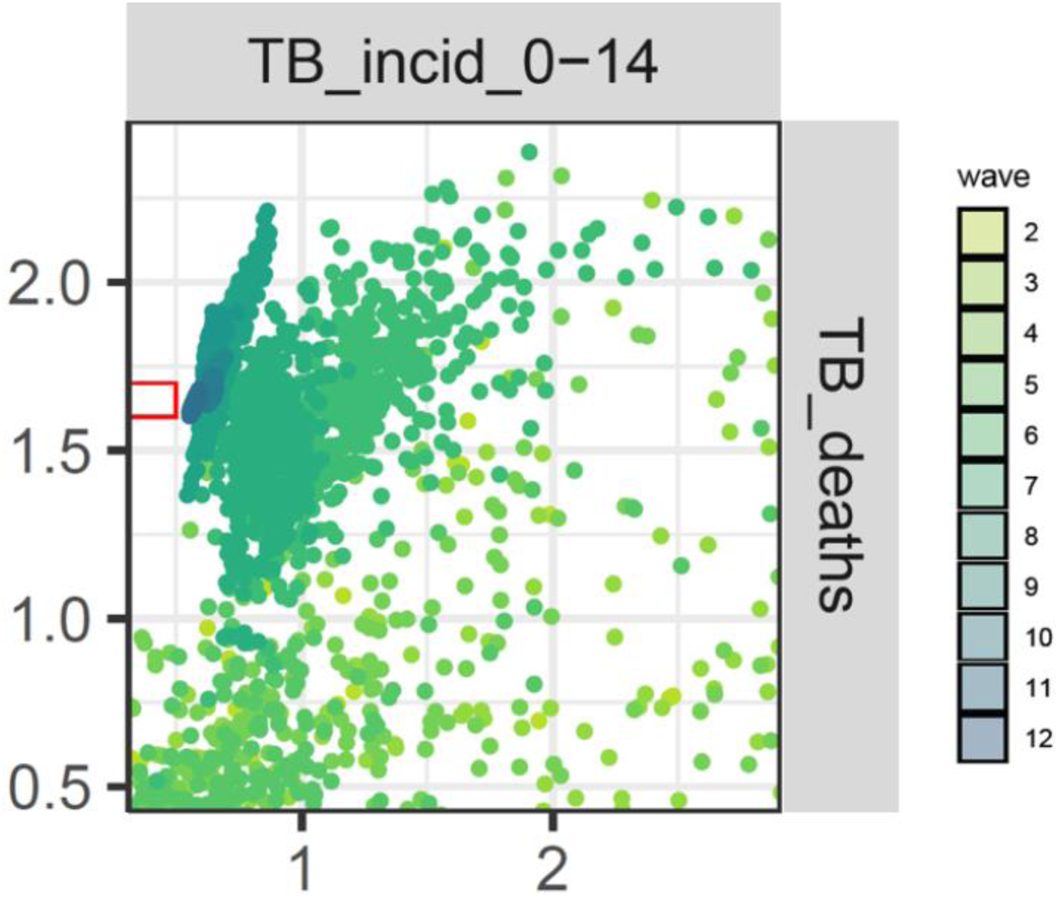
TB incidence in young individuals is possibly incompatible with TB deaths in all the population. Pair plot of TB incidence in young individuals and TB deaths, for parameter sets in waves 2, 3,…, 12 (country I).

## C. Bayes Linear Emulation

In the hmer package we adopt a Bayes Linear approach to build emulators. While a full Bayesian analysis requires specification of a full joint prior probability distribution to reflect beliefs about uncertain quantities, in the Bayes linear approach expectations are taken as a primitive and only first and second order specifications are needed when defining the prior. Operationally, this means that one just sets prior mean vectors and covariance matrices for the uncertain quantities, without having to decide exactly which distribution is responsible for the chosen mean and covariance. A Bayes Linear analysis may therefore be viewed as a pragmatic approach to a full Bayesian analysis, where the task of specifying beliefs has been simplified. As in any Bayesian approach, our priors (mean vectors and covariance matrices) are then adjusted by the observed data.

The general structure of a univariate emulator is as follows:

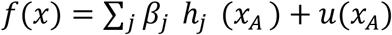

where ∑_*j*_ *β*_*j*_ *h*_*j*_ (*x*_*A*_) is a regression term and *u*(*x*_*A*_) is a weakly stationary process with mean zero. In the regression term, the sum is taken over the collection of regression functions *h*_*j*_ ′*s*, which can vary for different outputs. The argument of the *h*_*j*_ ′*s* and of the process *u* is *x*_*A*_ : this indicates the set of active variables for output *f*, i.e the variables that contributes the most to the value of *f*. The use of *x*_*A*_ instead of *x* as arguments of the *h*_*j*_ ′*s* and of *u* plays a key role in reducing the dimensionality of the problem, which is key when dealing with high-dimensional input spaces. The role of the regression term is to mimic the global behaviour of the model output, while the weakly stationary process represents localised deviations of the output from this global behaviour near *x*. In the regression term, the functions *h*_*j*_ ‘s determine the shape and complexity of the regression hypersurface we fit to the training data and the *β*_*j*_ ‘s are the regression coefficients. To fully describe the weakly stationary process *u*(*x*), we need to define the covariance structure, i.e. we need to say how correlated the local deviations at *x* and *x*′ are, for any pair (*x, x*′). The default option in the hmer package is to assume that *u*(*x*) is a Gaussian process with covariance structure given by

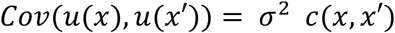

where *c* is the square-exponential correlation function

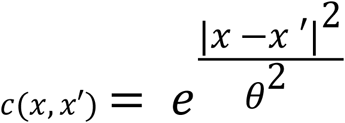

The multiplicative factor *σ*^2^ allows us to set a prior for the emulator variance, while *θ* is the correlation length, representing our belief about the smoothness of the emulated output (which can be generalised to an individual *θ*_*i*_ along each active input direction). The numerator |*x* − *x* ′|^2^in the exponent indicates the square of the distance between *x* and *x*′: Σ_*i*_ (*x*_*i*_ − *x*_*i*_ ′)^2^.

Once the structure of the emulator is chosen, prior mean vectors and covariance matrices are set, and a set of model runs *D* is available, we can train the emulator using the Bayes Linear Updates formulae:

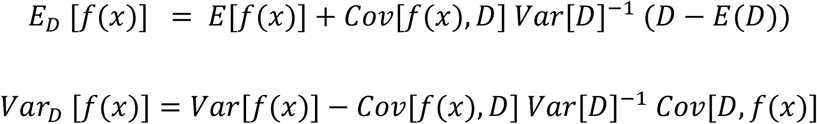

In these formulae, the expectations and variance on the right hand side refer to the set priors, while the left hand sides refer to the expectation and variance updated, based on the knowledge of the runs in *D*. Once an emulator has been built, it can be used to calculate the implausibility measure of any parameter set of interest. For a given model output *f* and a given target *z*, the implausibility is defined as the difference between the emulator output and the target, taking into account all sources of uncertainty:

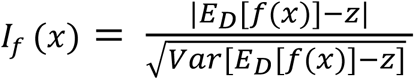

In this work, the variance in the denominator accounted for the emulator uncertainty *Var*_*D*_ [*f*(*x*)] and the observation uncertainty. Other forms of uncertainty can be included if relevant: for example, if dealing with a stochastic model, it is appropriate to factor in the uncertainty due to the ensemble variability of the model output.

## D. Reduction factor calculation

To quantify how much history matching with emulation reduced the input space, we first calculated the volume of the smallest hyper-rectangle containing all non-implausible points from the last wave of history matching for each of the countries and compared it to the volume of the original input space. Note that, especially when parameters are highly correlated, such a hyper-rectangle largely overestimates the size of the remaining space. To correct for this, we then accounted for the proportion of all points proposed at the end of the last wave that were non-implausible. This method estimated a median reduction factor of 6 × 10^8^, with 95% of all reduction factors lying in the interval [2 × 10^5^, 2.8 × 10^19^]. The median reduction factor per wave was 2.2 × 10^7^.

